# Quantifying absolute neutralization titers against SARS-CoV-2 by a standardized virus neutralization assay allows for cross-cohort comparisons of COVID-19 sera

**DOI:** 10.1101/2020.08.13.20157222

**Authors:** Kasopefoluwa Y. Oguntuyo, Christian S. Stevens, Chuan-Tien Hung, Satoshi Ikegame, Joshua A. Acklin, Shreyas S. Kowdle, Jillian C. Carmichael, Hsin-Ping Chiu, Kristopher D. Azarm, Griffin D. Haas, Fatima Amanat, Jéromine Klingler, Ian Baine, Suzanne Arinsburg, Juan C. Bandres, Mohammed N.A. Siddiquey, Robert M. Schilke, Matthew D. Woolard, Hongbo Zhang, COVIDAR Argentina Consortium, Andrew J. Duty, Thomas A. Kraus, Thomas M. Moran, Domenico Tortorella, Jean K. Lim, Andrea V. Gamarnik, Catarina E. Hioe, Susan Zolla-Pazner, Stanimir S. Ivanov, Jeremy P. Kamil, Florian Krammer, Benhur Lee

**Author notes:** These authors contributed equally to this work (Kasopefoluwa Y. Oguntuyo, Christian S. Stevens). COVIDAR Argentina Consortium: Ojeda DS, Gonzales Lopez Ledesma MM, Costa Navarro GS, Pallarés HM, Sanchez LN, Perez P, Ostrowski M, Villordo SM, Alvarez DE, Caramelo JJ, Carradori J, Yanovsky MJ, Gamarnik AV i. Fundacion Instituto Leloir-CONICET, Buenos Aires, Argentina ii. Universidad Nacional de San Martin, Buenos Aires, Argentina iii. Laboratorio Lemos SRL, Buenos Aires, Argentina iv. INBIRS-CONICET, Facultad de Medicina, Universidad de Buenos Aires, Argentina. Author contributions: KYO, CSS and BL conceived and designed the study. KYO, CSS, CTH, SI, JAA, SSK, JCC, HPC, KDA, GDH, FA, JK, IB, SA, JCB, SSI, MNAS, RMS, MDW, HZ, and COVIDAR Argentina Consortium collected data. GDH, JAD, TAK, TMM, DT, JKL, AVG, CEH, SZP, SSI, JPK, and FK contributed valuable reagents, data and/or tools. KYO and CSS analyzed the data and wrote the original drafts of the paper. BL reviewed the draft, supported data analysis, and provided invaluable direction throughout the conceptualization and execution of the project. All authors had the opportunity to review the manuscript prior to submission and SI, JCC, KDA, DT, and JPK provided valuable feedback during the editing process.

## Abstract

The global COVID-19 pandemic has mobilized efforts to develop vaccines and antibody-based therapeutics, including convalescent plasma therapy, that inhibit viral entry by inducing or transferring neutralizing antibodies (nAbs) against the SARS-CoV-2 spike glycoprotein (CoV2-S). However, rigorous efficacy testing requires extensive screening with live virus under onerous BSL3 conditions which limits high throughput screening of patient and vaccine sera. Myriad BSL-2 compatible surrogate virus neutralization assays (VNAs) have been developed to overcome this barrier. Yet, there is marked variability between VNAs and how their results are presented, making inter-group comparisons difficult. To address these limitations, we developed a standardized VNA using VSVΔG-based CoV-2-S pseudotyped particles (CoV2pp) that can be robustly produced at scale and generate accurate neutralizing titers within 18 hours post-infection. Our standardized CoV2pp VNA showed a strong positive correlation with CoV2-S ELISA and live virus neutralizations in confirmed convalescent patient sera. Three independent groups subsequently validated our standardized CoV2pp VNA (n>120). Our data show that absolute (abs) IC50, IC80, and IC90 values can be legitimately compared across diverse cohorts, highlight the substantial but consistent variability in neutralization potency across these cohorts, and support the use of absIC80 as a more meaningful metric for assessing the neutralization potency of vaccine or convalescent sera. Lastly, we used our CoV2pp in a screen to identify ultra-permissive 293T clones that stably express ACE2 or ACE2+TMPRSS2. When used in combination with our CoV2pp, we can now produce CoV2pp sufficient for 150,000 standardized VNA/week.

**Importance:** Vaccines and antibody-based therapeutics like convalescent plasma therapy are premised upon inducing or transferring neutralizing antibodies that inhibit SARS-CoV-2 entry into cells. Virus neutralization assays (VNAs) for measuring neutralizing antibody titers (NATs) is an essential part of determining vaccine or therapeutic efficacy. However, such efficacy testing is limited by the inherent dangers of working with the live virus, which requires specialized high-level biocontainment facilities. We therefore developed a standardized replication-defective pseudotyped particle system that mimics entry of live SARS-CoV-2. This tool allows for the safe and efficient measurement of NATs, determination of other forms of entry inhibition, and thorough investigation of virus entry mechanisms. Four independent labs across the globe validated our standardized VNA using diverse cohorts. We argue that a standardized and scalable assay is necessary for meaningful comparisons of the myriad of vaccines and antibody-based therapeutics becoming available. Our data provide generalizable metrics for assessing their efficacy.

## Introduction

Severe Acute Respiratory Syndrome Coronavirus 2 (SARS-CoV-2) is an enveloped, positive-sense, single-stranded RNA (+ssRNA) virus from the family *Coronaviridae*. SARS-CoV-2 is related to, but not derived from SARS-CoV, which we will refer to as SARS-CoV-1 for clarity. SARS-CoV-1 and SARS-CoV-2 belong to the genus *Betacoronavirus* and group together as sarbecoviruses, a subgenus that also contains numerous bat “SARS-like” CoVs.^1^ SARS-CoV-1 caused a limited epidemic of SARS from 2002-2004, infecting ~8,000 people and killing 774.^2,3^ SARS-CoV-1 was ultimately contained and has not reappeared. SARS-CoV-2 is the causative agent for coronavirus disease 2019 (COVID-19). The Chinese government first reported a cluster of 40 cases of atypical pneumonia (now known to be COVID-19) to the WHO on 30 Dec 2020. Since then, SARS-CoV-2 has erupted into a global pandemic, resulting in approximately 15 million cases and more than half a million deaths in less than 8 months.^4^

The emergence and spread of SARS-CoV-2 has required a global response to mitigate the fallout from the pandemic. As a result, the highest priorities for governments around the world are prevention, treatment, and monitoring of infection and immunity.^5^ Understanding and monitoring immune responses to SARS-CoV-2 is critical for development of antibody-based therapeutics and vaccines. Both are challenging to study at the necessary scale due to the inherent danger of working with live virus and limited access to high level biosafety containment facilities (i.e. BSL3). However, the development of pseudotyped viral particles capable of recapitulating SARS-CoV-2 entry—without the dangers or limitations of working with live virus—addresses these concerns. Many such pseudotype virus (PsV) systems based on lentivirus or vesicular stomatitis virus backbones have been published.^6–9^ These PsV systems have been used to understand and assess humoral immunity in acute and recovered COVID-19 patients, and to screen for therapeutic entry inhibitors, such as small molecules, monoclonal antibodies, or convalescent sera. Most importantly, such a surrogate BSL2 virus neutralization assay (VNA) is needed to screen for vaccine induced responses, in domestic animals and humans, as the world rushes to develop candidate vaccines against SARS-CoV2.

As of this writing, at least five SARS-CoV-2 vaccine developers have reported Phase I/II results involving over 1700 participants.^10–15^ While each group claims promising results, it is difficult to compare vaccine induced immune responses between the various vaccine platforms. This is not only due a lack of a standardized reporting but also due to a lack of standardized assays for reporting virus neutralization titers. Furthermore, at least 16 studies have reported 350 patients receiving convalescent plasma therapy for COVID-19. Across all 16 plasma studies, some groups establish enzyme-linked immunosorbent assays (ELISA) or live virus neutralization thresholds to screen donor plasma, while others do not report binding or neutralization data.^16-33^ Notably, none of these studies report using a PsV VNA to screen donor plasma. These discrepancies in screening methods/metrics limit the ability to compare across groups and make it difficult to draw conclusions about the quality/potency of antibody transferred to the recipient.^32,33^

A standardized virus neutralization assay (VNA) that provides robust, high-throughput results (>100,000 infections/week), is easily “kit-able”, and generates absolute virus neutralization titers (VNT), would allow for meaningful comparisons across different labs. In addition to helping down-select the myriad vaccine candidates, use of a standardized VNA to report VNT in absolute units can crowd-source the immense effort being expended by multiple labs across the globe to better understand the basis of the marked variation in VNT seen in COVID-19 recovered patients.^34,35^

The SARS-CoV-2 spike glycoprotein (S) is embedded in the viral envelope and facilitates both receptor recognition and membrane fusion. SARS-CoV-2-S is 1273 amino acids in length and, like other coronaviruses, is a trimeric class I fusion protein.^36^ The S glycoprotein contains two subunits, the N-terminal, S1 subunit and the C-terminal, S2 subunit. The S1 subunit contains the receptor-binding domain (RBD), which is responsible for host receptor binding. The S2 subunit contains the transmembrane domain, cytoplasmic tails, and machinery necessary for fusion, notably the fusion peptide and heptad repeats.^37,38^ Angiotensin-converting enzyme 2 (ACE2), a cell surface enzyme in a variety of tissues, facilitates binding and entry of SARS-CoV-2.^39–41^ However, ACE2 alone is not sufficient for efficient entry into cells. While entry depends on the S1 subunit binding ACE2, entry is further enhanced by proteolytic cleavage between the S1/S2 and S2’ subunits. For both SARS-CoV-1 and SARS-CoV-2, this cleavage-mediated activation of S-mediated entry is supported by the expression of cell-associated proteases, like cathepsins or transmembrane serine protease 2 (TMPRSS2), or the addition of exogenous proteases that mimic the various trypsin-like proteases present in the extracellular lung milieu.^39,42–51^ These proteases facilitate entry at the cell surface or via an endosomal route in a cell-type dependent manner. Extracellular proteases are thought to play a pathophysiogical role in the lung tissue damage caused by unabated MERS-CoV, SARS-CoV-1, and likely SARS-CoV-2 replication.^49,50^ Thus, in order to represent SARS-CoV-2 cell entry faithfully, a viral neutralization assay (VNA) must be sensitive not only to ACE2 binding but also to the proteolytic activation of spike.

In addition to its role in receptor binding and entry, S is the primary surface glycoprotein and is the major target of the neutralizing antibody response.^52–56^ Patients infected with SARS-CoV-2 typically seroconvert within two weeks of symptom onset, with about half developing antibodies within 7 days.^57–59^ Antibody titers appear to be durable at greater than 40 days post infection,^58^ but in the case of SARS-CoV-1, reductions in IgG positive titers begin around 4-5 months post infection and show a significant drop by 36 months.^60^ Although there are reports of SARS-CoV-2 infected individuals testing positive by RT-PCR weeks after being confirmed as recovered by two consecutive negative tests, these are more likely the result of false negatives than of reinfection.^61,62^ Multiple groups have shown that fully recovered rhesus macaques previously infected with SARS-CoV-2 are refractory to reinfection, at least within four weeks of the primary challenge.^63,64^ However, a better understanding of the durability and efficacy of the neutralizing antibody response in patients previously infected with SARS-CoV-2 is of paramount importance. Not only do IgG titers wane in the case of SARS-CoV-1, but reinfection is possible in other endemic human coronaviruses (HCoVs) such as 229E, NL63, and OC43 in as little as a year.^65–67^ Whether the waning of neutralizing SARS-CoV-2 antibodies impacts susceptibility to re-infection is an urgent question that needs to be answered by longitudinal follow-up studies.^68–71^

Humoral immune responses to the SARS-CoV-2 S protein are typically evaluated by ELISAs and its many variants (CLIA, LFA, etc.). These serological binding assays rightfully play a central role in determining patient antibody responses and can complement diagnostics and sero- epidemiological studies, especially when combined with antibody subclass determination (IgM, IgA and IgG).^72–74^ Nonetheless, as many antibodies generated to the spike protein bind but do not block virus entry,^75–78^ ELISA-based assays that detect titers of spike-binding antibodies cannot always correlate perfectly with neutralizing antibody titers as measured by plaque reduction neutralization or microneutralization tests.^74,79–82^ Even a cleverly designed competitive ELISA set up to detect antibodies that block the binding of RBD to ACE2^76,83^ cannot capture the universe of neutralizing antibodies targeted to a conformationally dynamic trimeric spike on a virion.^84,85^ The gold standard for detecting antiviral antibodies remains the virus neutralizing assay. Assays that faithfully recapitulate entry of SARS-CoV-2 while maximizing safety, speed, and scalability will be vital in the coming months and years. They will enable monitoring of patient neutralizing antibody response, efficacy of vaccines and entry inhibitors, and the screening of convalescent plasma from COVID-19 recovered patients.^57,86^

In order to meet this need while maximizing safety, speed, and scalability, we generated a SARS-CoV-2 pseudotyped viral particle (CoV2pp) by using vesicular stomatitis virus bearing the *Renilla* luciferase gene in place of its G glycoprotein (VSVΔG-rLuc). This approach has been used safely by our group and others to study viruses that would otherwise require significant biosafety constraints, including Ebola virus, Nipah virus, and, most recently, SARS-CoV-2.^6,8,87–90^ Here, we present a detailed protocol for the production of CoV2pp, characterize the contributions of stable expression of ACE2 as well as endogenous or exogenous proteases on entry, and standardize the production and performance characteristics of these CoV2pp for use in a robust high throughput VNA. We have sent out our standardized CoV2pp as ready-to-use “out of the box” VNAs, ≥1000 infections/request, to multiple labs across three continents. We show here the validation of our CoV2pp in a standardized VNA by four independent groups spread across two continents using sera samples from geographically distinct and ethnically diverse cohorts. Lastly, we utilized our standardized CoV2pp and VSV-Gpp in a screen to identify two ultra-permissive 293T cell clones that stably express either ACE2 alone or ACE2+TMPRSS2. These isogenic cell lines support either the late (293T-ACE2) or early (293T-ACE2/TMPRSS2) entry pathways that SARS-CoV-2 uses.^40,45,50,91^ These ultra-permissive 293T clones allow for use of unpurified virus supernatant from our standard virus production batch, which can now provide for ~150,000 infections per week (96-well format) with no further scale-up. In sum, we have generated a standardized, scalable, high-throughput BSL2-compatible CoV2pp VNA that can provide robust metrics (absIC50, absIC80, absIC90) for meaningful comparisons between labs.

## Results

### Production of VSVAG-rLuc bearing SARS-CoV-2 spike glycoprotein

Our initial objective was to produce SARS-CoV-2 PsV sufficient for ≥10,000 infections/week at ~1:100 signal:noise ratio when performed in a 96-well format. We settled on a VSV-based rather than a lentiviral PsV system as lentiviruses are intrinsically limited by their replication kinetics and particle production rate (10^4^-10^6^/ml for lentiviruses versus 10^7^-10^9^/ml for VSV without concentration). We optimized the production of our VSVΔG-rLuc pseudotyped viral particles (pp) bearing the SARS-CoV-2 spike glycoprotein as diagramed in Figure 1A. A detailed production protocol is given in Supplementary Methods. Notably, this protocol involves infecting producer cells at a low multiplicity of infection (MOI) of stock VSVΔG-G*, incubating producer cells with an anti-VSV-G monoclonal antibody and generating the pseudotyped particles in Opti-MEM media. The first two measures effectively eliminated the background signal from residual VSV-G while the last measure allowed for more cleavage of SARS-CoV-2pp in producer cells (Supplemental Figure 1). While others have shown that truncating the cytoplasmic tail (CT) of SARS-CoV-2-S is typically required for greater functional incorporation into heterologous viral cores, ^7,9,92,93^ we chose to optimize pseudotyping with full-length SARS-CoV-2 spike. CT truncations in many other class I viral fusion proteins, including other ACE2-using coronaviruses (HCoV-NL63 and SARS-CoV-1) can affect ectodomain conformation and function.^94–103^ Until such time that we gain a fuller understanding of SARS-CoV-2 entry, we felt it was necessary to have a surrogate assay that reflects the biology of the full-length virus spike.

**Figure 1.**
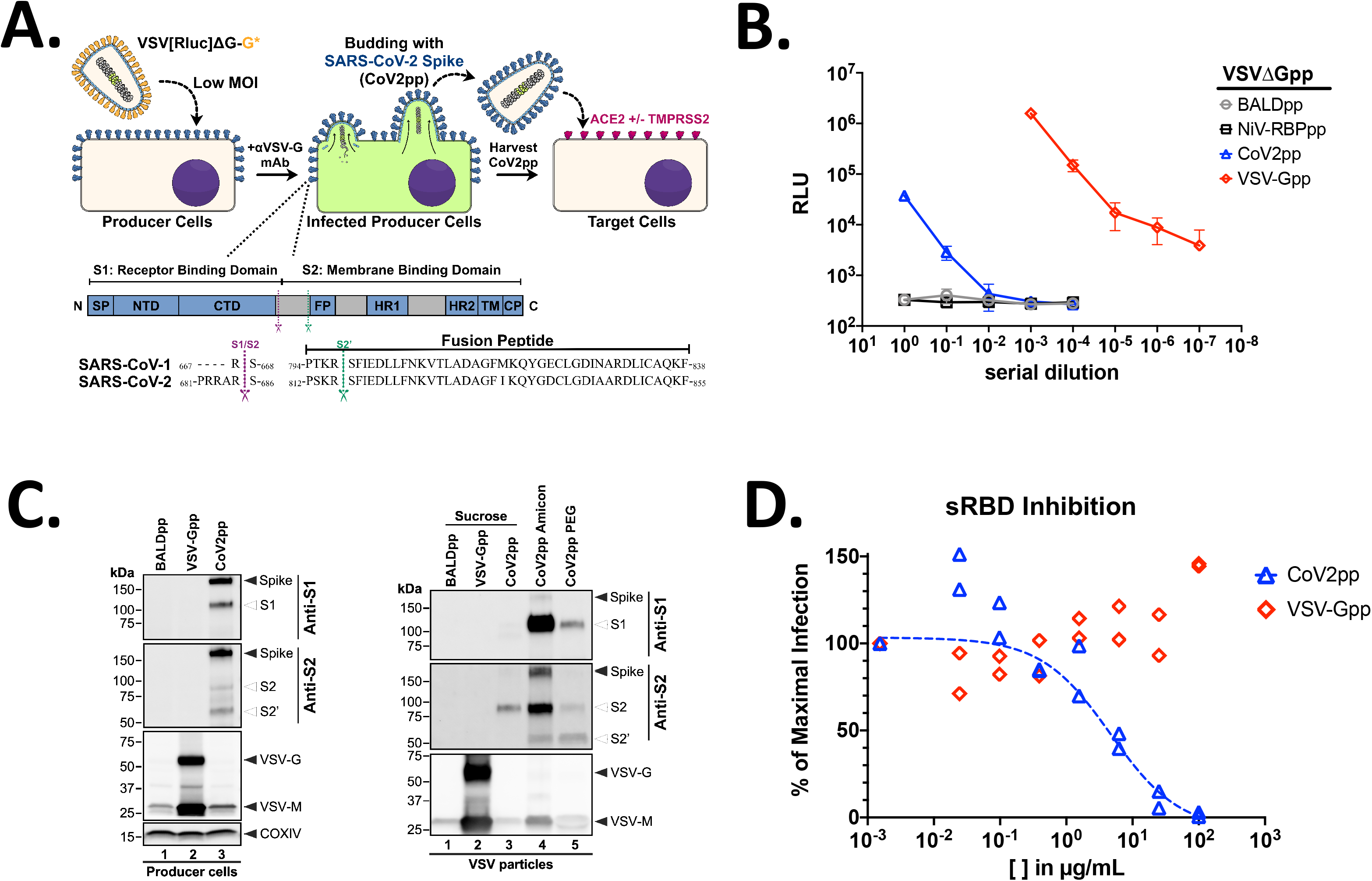
Production of VSVΔG-rLuc bearing SARS-CoV-2 spike glycoprotein. **(A)** Overview of VSVΔG-rLuc pseudotyped particles bearing CoV-2 spike (top panel) with annotated spike glycoprotein domains and cleavage sites (bottom panel). As mentioned in the text, we refer to SARS-CoV as SARS-CoV-1 for greater clarity. **(B)** VSV-AG[Rluc] pseudotyped particles (VSVpp) bearing the Nipah virus receptor binding protein alone (NiV- RBPpp), SARS-CoV-2-S (CoV2pp), or VSV-G (VSV-Gpp) were titered on Vero-CCL81 cells using a 10-fold serial dilution. Symbols represent the mean +/- SEM (error bars) of each titration performed in technical triplicates. **(C)** Expression of the indicated viral glycoproteins on producer cells and their incorporation into VSVpp. Western blots performed as described in Methods using anti-S1 or anti-S2 specific antibodies. **(D)** CoV2pp entry is inhibited by soluble receptor binding domain (sRBD) derived from SARS-CoV-2-S. CoV2pp and VSV-Gpp infection of Vero-CCL81 cells was performed as in (B) in the presence of the indicated amounts of sRBD. Neutralization curves were generated by fitting data points using a variable slope, 4-parameter logistics regression curve (robust fitting method). The last point (no sRBD) was fixed to represent 100% maximal infection. Each replicate from an experiment performed in duplicate is shown. The calculated IC50 for sRBD neutralization of CoV2pp is 4.65μg/mL.

Following the protocol detailed in Supplementary Methods, we produced BALDpp, NiV-RBPpp, CoV2pp, and VSV-Gpp using the VSVΔG-rLuc reporter backbone and titered them on Vero-CCL81 cells (Fig. 1B). High background problems have resulted in low signal:noise ratios when using VSV-based PsV, especially for viral envelope proteins that do not mediate efficient entry. Here we used two different negative controls, BALDpp and NiV-RBP, to show that we resolved the background issue. BALDpp lacks any surface glycoprotein while NiV-RBPpp incorporates the NiV receptor binding protein (RBP), which binds to the broadly expressed ephrin-B2 with sub-nanomolar affinity.^88,104^ However, the NiV fusion (F) glycoprotein necessary for viral entry is absent. NiV-RBPpp without NiV-F should not fuse and effectively serves as a stricter and complementary negative control. Under the conditions shown, neither BALDpp or NiV-RBPpp gives any background even at the highest concentration of virus particles used.

These constructs were used to infect Vero-CCL81 cells and, as expected, we observe an average of <500 RLUs of entry with our BALDpp and NiV-RBPpp negative controls. These levels of entry were comparable to the “cells only” signal, providing confidence in any infection signals 10-fold over background. Undiluted CoV2pp entry resulted in luciferase values of over 50,000 RLUs; greater than 100-fold over background BALDpp signals (Fig. 1B). VSV-Gpp gave several logs higher infectivity as expected. Western blots of the producer cells demonstrated effective expression of cleaved, SARS-CoV-2 spike glycoproteins (Fig. 1C, left panel). Cleaved CoV-2 spike products (S1, S2, and S2’) all appear to be incorporated into the VSVAG pseudotyped particles (Fig. 1C, right panel). To ensure that entry of CoV2pp is SARS-CoV-2 spike-mediated, we show that the homologous soluble spike receptor binding domain (sRBD) competitively inhibits our CoV2pp (Fig. 1D).

### CoV2pp entry is enhanced by trypsin treatment and spinoculation

Next, we sought to enhance the relative signal of our CoV2pp infections, which will effectively increase the number of infections we can provide or perform per batch of CoV2pp. Trypsin treatment is reported to enhance SARS-CoV-1 and SARS-CoV-2 entry.^39,45^ Thus, we treated CoV2pp stocks with the indicated range of trypsin concentrations for 15 min at room temperature (Fig. 2A). In order to mitigate the effects of trypsin-dependent cytotoxicity, we added 625μg/mL of soybean trypsin inhibitor (SBTI) to all samples before titrating the trypsin-treated CoV2pp onto Vero-CCL81 cells. CoV2pp treated with the highest concentration of trypsin (625μg/mL) resulted in ~100-fold enhancement of entry (Fig. 2A), but this trypsin-dependent enhancement was only apparent when comparing entry of undiluted trypsin-treated CoV2pp. We observed a greater than 50-fold reduction in entry (RLUs) after a 10-fold serial dilution, which nullified any entry enhancement effects of trypsin. Indeed, the role of trypsin in enhancing SARS-CoV-2 entry has not been fully determined. Trypsin may be acting to prime CoV2pp to facilitate better entry upon spike-receptor interactions and/or assist to proteolytically activate spike protein at or after receptor binding.^50^ We hypothesized that the remaining uninhibited trypsin-dependent effect, which must be present at the highest trypsin concentration, was inadvertently neutralized by diluting the trypsin-treated CoV2pp in Dulbecco’s modified Eagle Medium (DMEM) +10% fetal bovine serum (FBS), which is the standard infection media for titrating CoV2pp. To test this hypothesis, we diluted CoV2pp and trypsin-treated CoV2pp 1:10 in three different media conditions before infecting Vero-CCL81 cells. For trypsin-treated CoV2pp, dilution in DMEM alone (serum free media, SFM) produced the highest signal:noise ratio, almost 1000-fold over BALDpp (Fig. 2B). As a result, we chose CoV2pp treated with 625μg/mL of TPCK-treated trypsin, then 625μg/mL of SBTI, diluted in SFM as our standard treatment condition. Furthermore, spinoculation at 1,250rpm for 1hr enhanced entry 3-5 fold (compare signal:noise in Fig. 2B to Supplemental Fig. 2).

**Figure 2.**
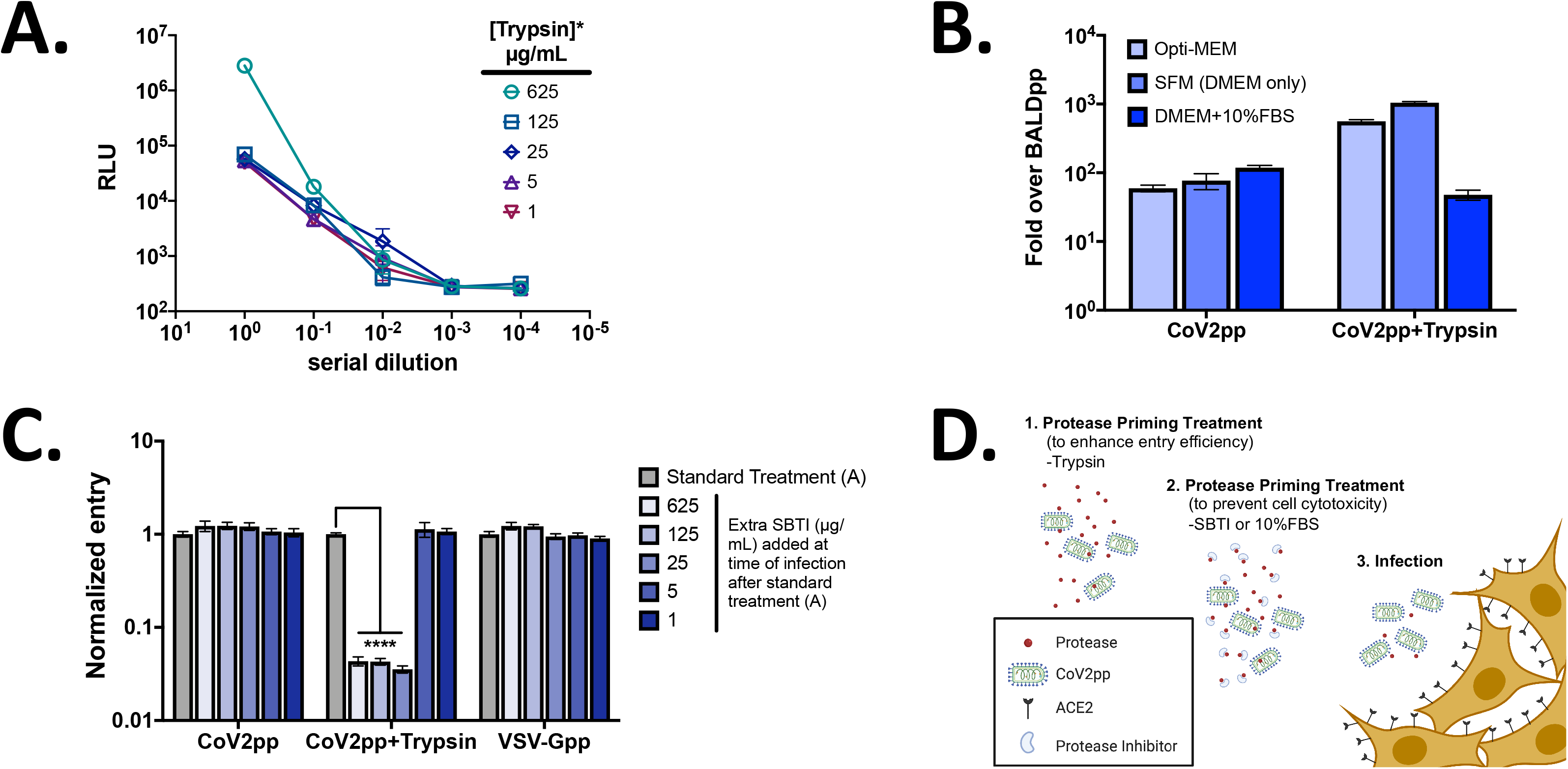
CoV2pp entry is enhanced by trypsin treatment. **(A)** Optimizing trypsin treatment conditions. Supernatant containing CoV2pp were trypsin-treated at the indicated concentrations for 15 min. at room temperature prior to the addition of 625 μg/mL of soybean trypsin inhibitor (SBTI). These particles were then titered on Vero-CCL81 cells in technical triplicates. Data shown as mean +/- SEM. **(B)** Dilution in serum free media (SFM, DMEM only) provides the highest signal:noise ratio for trypsin-treated CoV2pp entry. Particles were diluted 1:10 in Opti-MEM, SFM, or DMEM+10%FBS prior to infection of Vero-CCL81 cells and spinoculation as described in Fig. 1D. Cells infected without spinoculation show approximately 3x less signal:noise ratios (Supplemental Fig. 2). **(C)** Addition of soybean trypsin inhibitor at the time of infection reduces trypsin treated particle entry. This was performed in technical triplicates for two independent experiments. Shown are the combined results with error bars indicating SEM and **** indicating a p-value <0.0001. (D) Schematic showing overall view of how protease priming and SBTI treatment is working to enhance CoV2pp entry.

Our above hypothesis suggests that the uninhibited trypsin-dependent enhancing effect was acting at the point of infection when CoV2pp is interacting with the host cell receptor. To investigate further, we spiked in additional SBTI onto cells at the time of infection using particles produced under the standard treatment condition as above. We found that additional SBTI (≥25μg/mL) added directly to cells at the point of infection was able to inhibit trypsin-dependent entry enhancement (Fig. 2C). The data suggest that some trypsin was not inhibited by the first 625μg/mL of SBTI and enough remained to enhance entry at the point of infection (Fig. 2D).

### Entry of CoV2pp is independently enhanced by stable expression of ACE2 and TMPRSS2 in cells already permissive for SARS-CoV-2 entry and replication

To further characterize the determinants of CoV2pp entry, we generated Vero-CCL81 cell lines stably expressing human ACE2 or human TMPRSS2. Vero-CCL81 cells are already highly permissive for SARS-CoV-2 entry and replication. We infected the indicated cells with CoV2pp or trypsin-treated CoV2pp diluted in serum-free media (standard treatment) and observed enhanced entry in both stable cell lines (Fig. 3A). However, the entry enhancement of trypsin-treated CoV2pp in Vero-CCL81+TMPRSS2 overexpressing cells was subdued relative to untreated CoV2pp. This suggests that the presence of exogenous trypsin during CoV2pp entry can substitute, in part, for the role played by cell surface TMPRSS2, an endogenous protease known to facilitate entry into physiological relevant cell types *in* vivo.^105^ Fig. 3B shows that the relationship between ACE2 and TMPRSS2 expression—with regard to their effect on enhancing SARS-CoV-2 spike mediated entry—is not straightforward. As ACE2 itself is a substrate for TMPRSS2, the right stoichiometry of receptor/protease expression appears to be the main driver of entry efficiency rather than the absolute expression of one or the other. This issue will be further examined in the last section.

**Figure 3.**
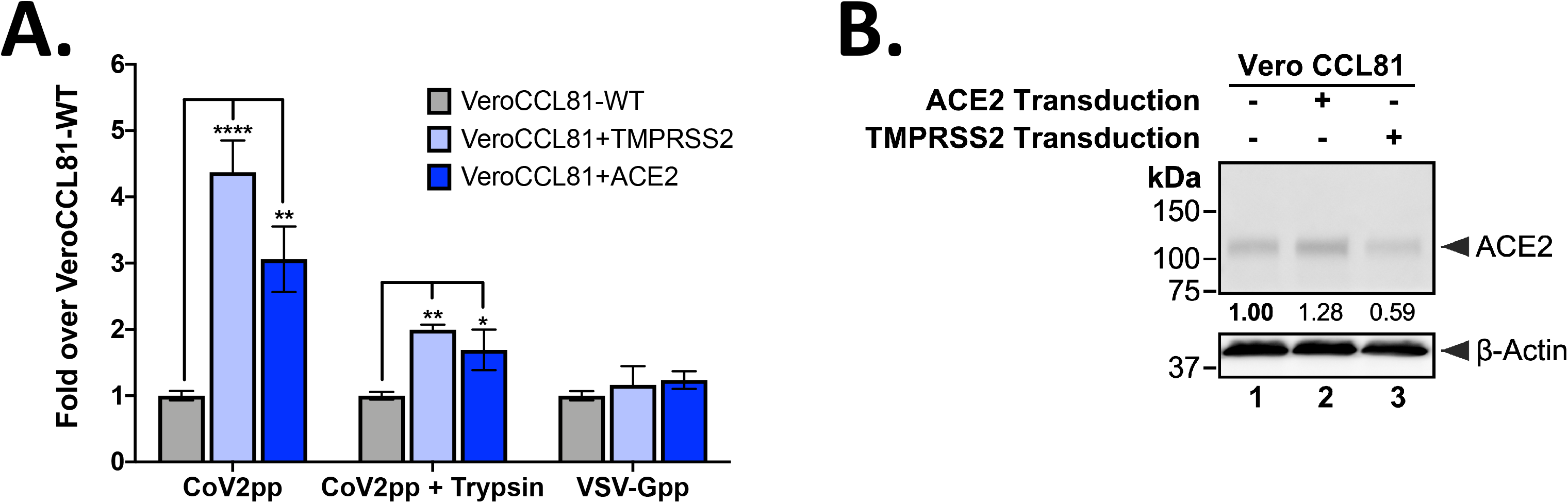
Trypsin-treated CoV2pp depend on ACE2 and TMPRSS2 for entry. **(A)** Parental and TMPRSS2 or ACE2 transduced VeroCCL81 cells were infected with the indicated pseudotyped viruses. All particles were diluted in serum free media in order to be within the linear range for the assay. Normalized infectivity data is presented as fold-over Vero-CCL81-WT for the various VSVpp shown. VSV-Gpp served as an internal control for the intrinsic permissiveness of various cell lines to VSV mediated gene expression. Data is presented as mean +/- SEM from two independent experiments done in technical triplicates. *, p <0.05, **, p-<0.01, and ****, p <0.0001. **(B)** Western blot of wild type and transduced Vero CCL81 cells. The numbers below each column show the relative ACE2 abundance was measured by densitometry and normalized as described in Methods.

### Standardizing the parameters that impact CoV2pp-based virus neutralization assay

Having established that exogeneous trypsin can serve as a physiologically relevant substitute for endogenous proteases known to enhance entry of CoV2pp, such as TMPRSS2, we sought to characterize the parameters that might affect the performance our CoV2pp VNA. Conditions tested included heat-inactivation of sera and the infection media used to dilute human sera samples. We used representative spike ELISA positive or negative sera to serve as positive and negative controls, respectively. When first diluted in SFM, we observed that negative sera can have alarming amounts of neutralizing activity that appeared specific for CoV2pp as the same sera did not neutralize VSV-Gpp entry (compare Supplemental Figures 3A with 3B, right panel). This CoV2pp serum neutralizing factor is somewhat reduced but not completely diminished by heat inactivation for 1hr at 56°C. Notably, the effect of this neutralizing factor from negative sera was preempted by diluting the trypsin treated CoV2pp in DMEM containing 10% FBS (Supplemental Fig. 3B). Importantly, recombinant sRBD neutralization was not affected by the dilution of CoV2pp in Serum Free Media or DMEM+10% FBS (Supplemental Fig. 3C). The nature of this factor that appears to inhibit spike-mediated entry is the subject of a concurrent manuscript in submission (see Discussion). Regardless, for standardizing our CoV2pp-based VNA, all subsequent patient sera were heat inactivated for at least 30 mins prior to use and serially diluted in DMEM + 10% FBS, which also served as our infection media. Despite our data from Fig. 2 implicating a trypsin-inhibitor-like activity in FBS, the marked inhibition of CoV2pp entry by seronegative human sera is a greater limiting factor that prevents the robust determination of true SARS-CoV-2 Nab titers. To achieve the same signal:noise ratio while performing our VNA in the presence of 10% FBS, we increased the concentration of CoV2pp used per infection.

### Performance characteristics of our standardized CoV2pp virus neutralization assay

An initial set of sera for validation of CoV2pp VNA was generously provided by Dr. Florian Krammer. These sera were screened according to a previously described two-stage ELISA protocol in which 1:50 dilutions of patient sera were first screened for reactivity against sRBD. Subsequently, the presumptive RBD-positive patient sera were used to assess reactivity to the trimer stabilized ectodomain of spike at five different dilutions (1:80, 160, 320, 960, and 2880).^73,106^ These samples were used for neutralization studies with CoV2pp (Fig. 4A and 4B). From the 36 patient sera tested, 6 were found to be negative for SARS-CoV-2 spike binding in the ELISA described above. All of those 6 sera samples also showed no neutralization of CoV2pp. The remaining 30 spike positive sera had 50% neutralizing titers that span 2 orders of magnitude (Fig. 4B, 160 – 10,240). For a more quantitative assessment, we determined the total IgG and IgM spike binding activity (ELISA AUC as described in Methods) of a representative subset of fifteen sera samples and compared them with their reciprocal absIC50 and absIC80 values calculated from the CoV2pp neutralization curves (Fig.4A) as described in Methods.

**Figure 4.**
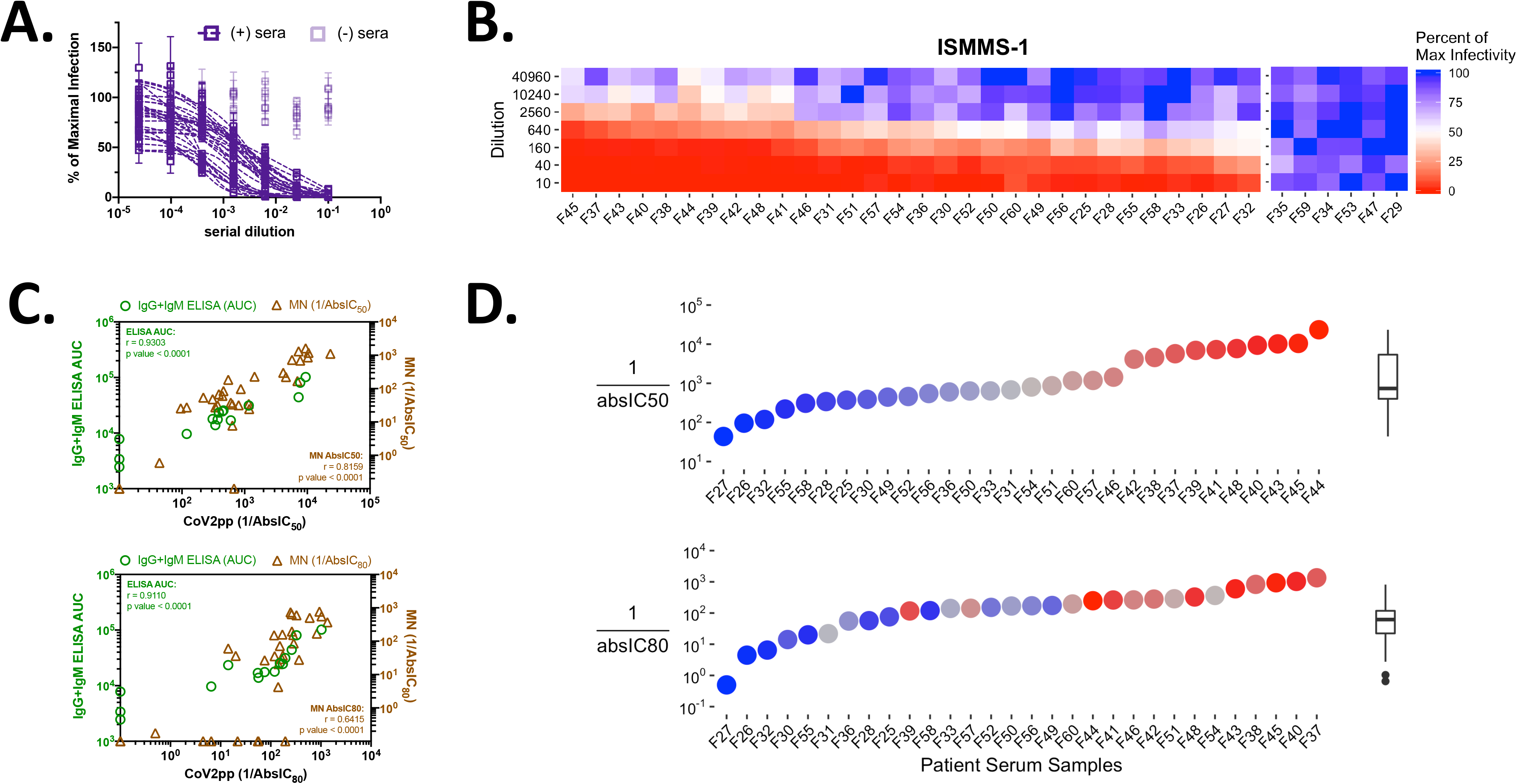
CoV2pp viral neutralization assay and absIC50/80 versus spike binding of patient sera. **(A)** 36 patient sera screened for CoV2pp neutralization. CoV2pp were used to infect Vero-CCL81 cells in the presence of a 4-fold serial dilution of patient sera as described in the Methods. Samples in light purple do not neutralize CoV2pp. Neutralization curves were fit using a variable slope, 4-parameter logistics regression curve with a robust fitting method. **(B)** The same 36 samples are shown as a neutralization heat map, which were generated in R as described in the Methods. Here, red represents complete neutralization and blue represents no neutralization. Samples are sorted by the average from the first four dilutions with the most neutralizing samples on the left. **(C)** Correlation of CoV2pp neutralization titers to spike binding (ELISA AUC) and live virus microneutralization (MN) activity. Absolute IC50 (absIC50, top) and IC80 (absIC80, bottom) for CoV2pp neutralizations and live virus MNs were calculated in R using a 4-parameter logistic regression model as described in the Methods. Presented are the added IgG and IgM ELISA AUC. AUC and live virus neutralizations were performed as described in the Methods. Presented are the r and p value from a simple linear regression. **(D)** Positive serum samples and their CoV2pp reciprocal absIC50 (top) and absIC80 (bottom). The IC50 graph is colored and ordered to display samples with low, average, or high IC50 as blue, grey or red circles, respectively. The IC80 graph below retains the coloring from the IC50 graph, but the samples are now ordered from left to right to show samples with the lowest to highest IC80 values. Tukey box and whisker plots show median with interquartile range (IQR) and whiskers extending to 1.5x the IQR. All points outside that range are depicted.

Spike binding antibodies (IgG+IgM ELISA AUC) demonstrated a significant, positive correlation with neutralizing antibody (nAb) titers (reciprocal absIC50 and absIC80) as determined by our CoV2pp VNA (Fig. 4C, green circles). Moreover, these Nab titers against CoV2pp also correlated well with live virus microneutralization titers (MN absIC50, MN absIC80) (Fig. 4C, brown triangles). Full neutralization curves for the MN titers are shown in Supplemental Figure 4. AbsIC80 appeared to be a more stringent measure of nAb activity, as some sera that have respectable MN absIC50 titers never achieve an absIC80 (Fig. 4C, bottom graph, brown triangles on the x-axis). In this respect, the CoV2pp VNA has a larger dynamic range and was more sensitive in its ability to sort out sera samples that can reach their respective absIC80 values. Notably, we find that sera samples with potent absIC50 titers do not always display potent absIC80 values (Fig. 4D).

### Independent validation of our CoV2pp VNA with geographically distinct and ethnically diverse COVID-19 patient cohorts

To assess the robustness of our standardized CoV2pp VNA, we produced and distributed the CoV2pp to many labs who have requested our assay for use in various screens for nAbs. Here, we analyze and present the raw virus neutralization data provided to us by three independent groups at the Icahn School of Medicine at Mount Sinai (ISMMS-2), Louisiana State University Health Sciences Center Shreveport (LSUHS), and Argentina (COVIDAR). In sera or plasma neutralization studies, these groups also observe similar absIC50, absIC80, and absIC90 distributions. The LSUHS and ISMMS-2 cohorts represent data from 25 and 28 seropositive as well as 10 and 11 seronegative samples, respectively, while the COVIDAR consortium assessed neutralization from an initial set of 13 seropositive patient samples. For clarity, analysis of their neutralization curves is presented as heatmaps in Fig. 5A similar to what was shown in Fig. 4B. Full neutralization curves for each cohort are shown in Supplemental Figure 5.

The seronegative control samples from all groups revealed no CoV2pp neutralization. Rare, but notable, seropositive samples from LSUHS also showed no neutralization (Fig. 5A, LSUHS). ISMMS-2 performed their analysis on confirmed convalescent plasma donors.^32^ While all donors had detectable nAb titers, their titers were highly variable and ranged across 2-3 logs. AbsIC80s were calculated for all samples shown and we observed a moderate, but significant, positive correlation between various spike ELISA metrics and absIC80 (Fig. 5B).

**Figure 5.**
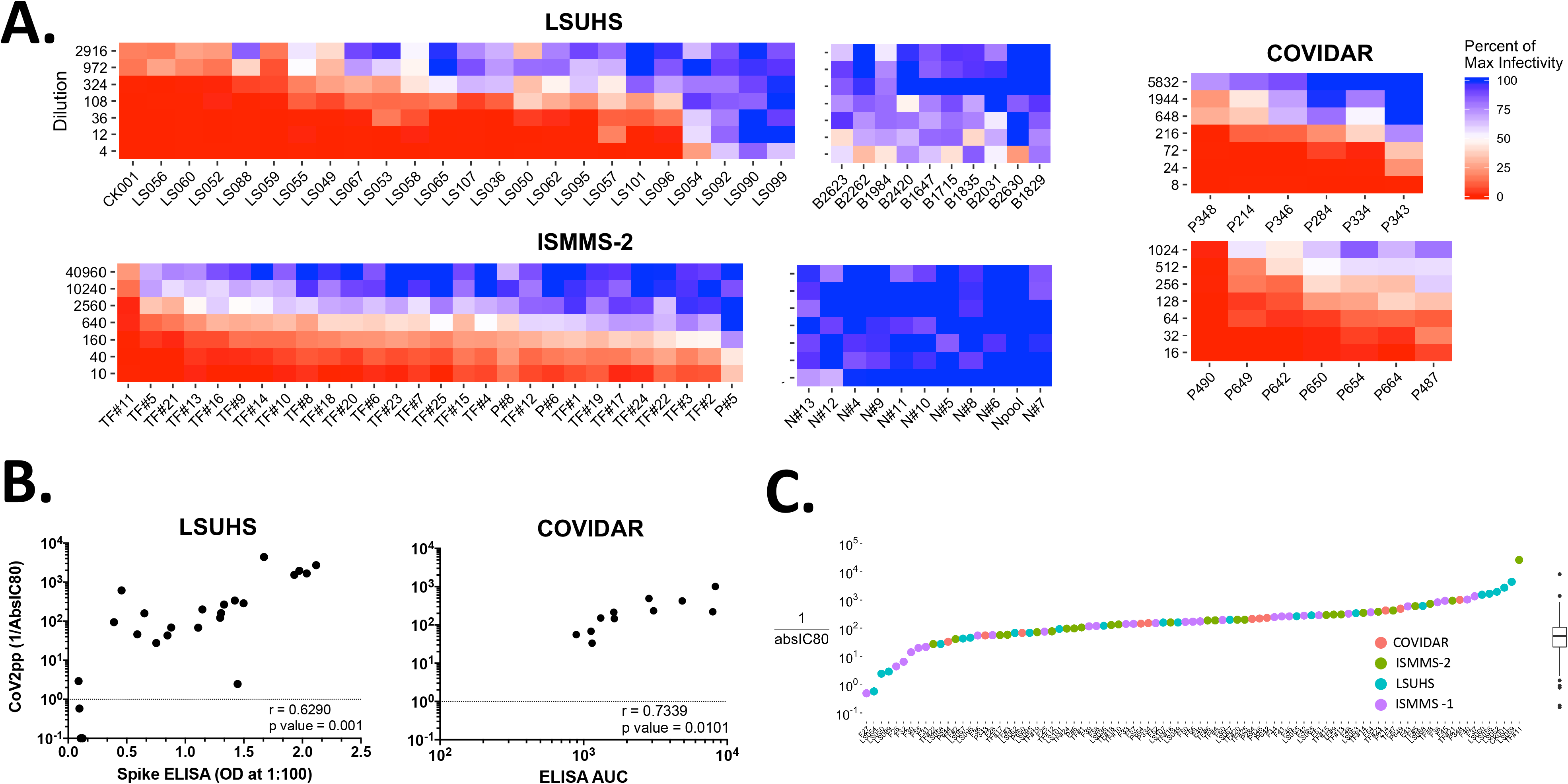
CoV2pp viral neutralization assay validated against patient sera by external groups. **(A)** Patient sera neutralization of CoV2pp for 88 samples run by three different independent groups. This is visualized as in Fig. 4B where red represents complete neutralization and blue represents no neutralization. **(B)** Correlations of CoV2pp reciprocal AbsIC80 to spike ELISAs. AbsIC80 was calculated as previously described in Fig. 4C. For LSUHS ELISAs, spike ectodomain was used and sera was diluted to a 1:100 dilution. For the COVIDAR ELISAs, a mixture of sRBD and spike was utilized as previously described^114^ and AUC was calculated as described in the Methods. **(C)** Summary AbsIC80 of 89 positive sera CoV2pp neutralizations. Samples from all 4 groups are depicted on the X-axis. AbsIC80 was calculated as described in Fig. 4C and Tukey box and whisker plots are shown as described in Fig. 4D.

Aggregated reciprocal absIC80 from all three external labs as well as our own are shown in Fig. 5C. Notably, we observe a Gaussian distribution of reciprocal absIC80s from all groups (n=89). The descriptive statistics from this aggregated data set reveals reciprocal absIC80 25^th^ percentile of 68.5, median of 170.8, and 75^th^ percentile of 343.4. Descriptive statistics for reciprocal absIC50 and absIC90 were also calculated and are reported in Supplemental Table 1. Using the absIC80 descriptive statistics above and the ELISA endpoint titers from our initial 36 sera samples, we observe that 0% of the samples displaying an ELISA endpoint titer of 320 have an absIC50 greater than the median IC50. Perhaps not surprisingly, over 90% of samples with ELISA endpoints of 2880 have IC50s at or beyond the 75th percentile (Table 1, represented graphically in Supplemental Figure 6). Although absIC80 also generally follows this trend, we once again note differences in the ranked order of absIC50 and absIC80 values calculated for all sera samples (Supplemental Figure 7). This difference is more pronounced when comparing the absIC50 and absIC90 graphs further highlighting the need for a neutralization assay with a broad dynamic range. Additionally, the samples from each of the 4 groups show no statistical difference when absIC50, 80, or 90 calculations are compared (Supplemental Figure 8). Altogether, these data support the robustness of our CoV2pp VNA and suggest that absIC80 is a more stringent and meaningful measure of Nab titers.

**Table 1.** Comparison of ELISA endpoint titers to CoV2pp neutralization. Presented are the clinical lab ELISA endpoint titers from samples discussed in Fig. 4A. Descriptive statistics were generated in PRISM using data presented in Fig. 5C. Values highlighted in red are of interest and are discussed further in the Results.

|  | IC<sub>50</sub> Summary [fraction of samples (%)] |  |  | IC<sub>80</sub> Summary [fraction of samples (%)] |  |  |
| --- | --- | --- | --- | --- | --- | --- |
|  | ≥ 25<sup>th</sup> percentile | ≥ median | ≥ 75<sup>th</sup> percentile | ≥ 25<sup>th</sup> percentile | ≥ median | ≥ 75<sup>th</sup> percentile |
| 320 | 7/11 (63.6%) | 0/11 (0%) | 0/11 (0%) | 4/11 (36.4%) | 1/11 (9.1%) | 0/11 (0%) |
| 960 | 6/8 (75%) | 4/8 (50%) | 0/8 (0%) | 7/8 (87.5%) | 4/8 (50%) | 1/8 (12.5%) |
| 2880 | 11/11 (100%) | 11/11 (100%) | 10/11 (90.9%) | 11/11 (100%) | 10/11 (90.9%) | 5/11 (45.5%) |

### Ultra-permissive 293T-ACE2 and 293T-ACE/TMPRSS2 clones allow for use of CoV2pp in VNA at scale

Although our standardized VNA appears robust, the requirement for exogenous trypsin and spinoculation to achieve the optimal signal:noise limits the scalability of our VNA. Therefore, we used our untreated CoV2pp to screen for ultrapermissive cell lines that would allow for our CoV2pp VNA to be performed with dilutions of virus supernatant without any trypsin treatment, virus purification, or spinoculation.

We generated three different 293T cell lines stably expressing ACE2 and/or TMPRSS2 via lentiviral transduction. We then infected these cells with CoV2pp. Increased expression of TMPRSS2 alone (293T-TMPRSS2) did not significantly improve entry (Fig. 6A), likely due to the low to undetectable ACE2 expression levels (Fig. 6B, lanes 1 and 3). However, expression of ACE2 significantly increased the entry of CoV2pp, which was further increased in 293T-ACE2+TMPRSS2 cells, suggesting the synergistic activity of TMPRSS2 and ACE2 (Fig. 6A). Western blot analysis confirmed the increased expression of ACE2 in the singly and doubly transduced 293T cells (Fig. 6B). Additionally, increased expression of both ACE2 and TMPRSS2 was confirmed by qPCR (Supplemental Fig. 9B). Interestingly, ACE2 expression appeared to be decreased by >50% in 293T-ACE2+TMPRSS2 cells relative to 293T-ACE2 cells.

**Figure 6.**
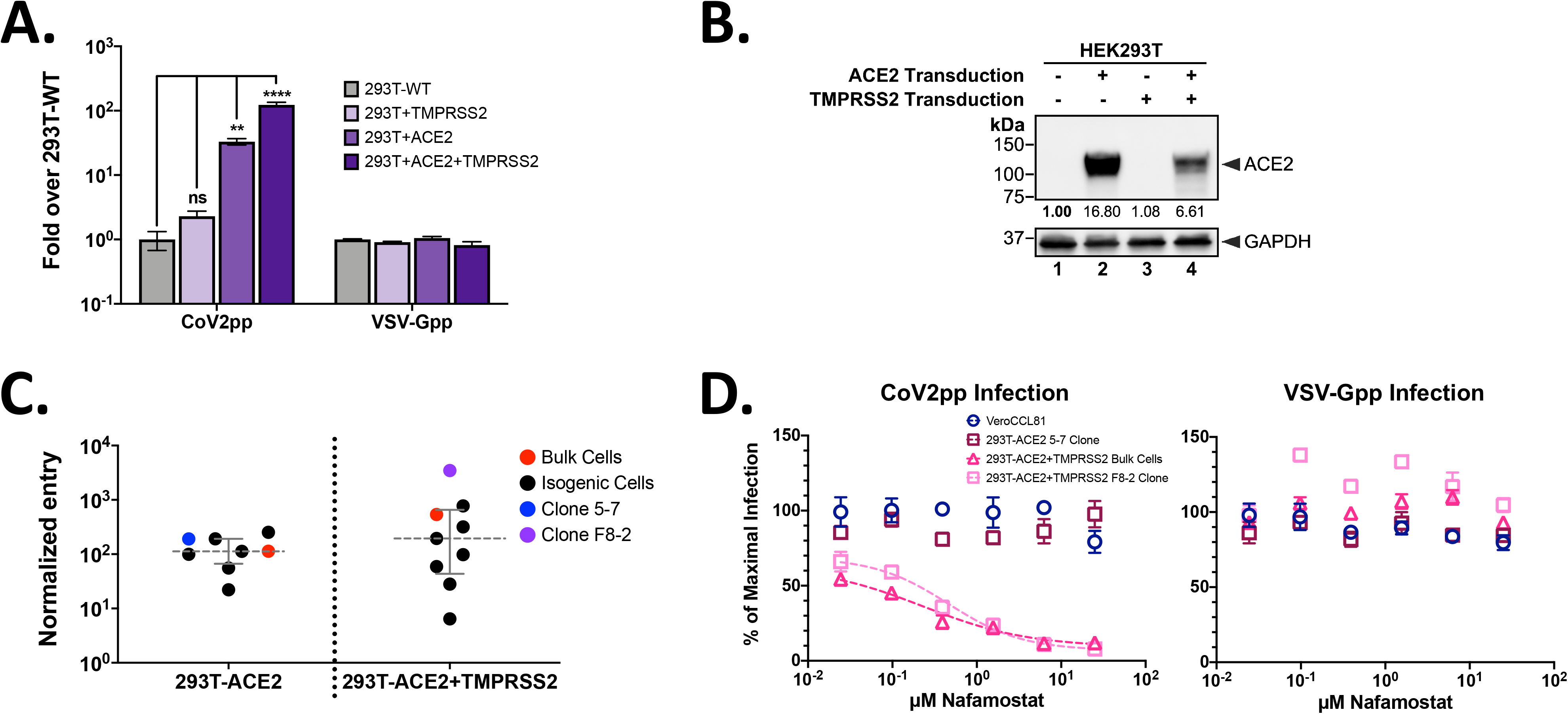
293T stably transduced with ACE2 and TMPRSS2 (293T-ACE2+TMPRSS2) are ultra-permissive for SARS-CoV-2pp infection. **(A)** Infection of 293T cells lines transduced to stably express, TMPRSS2, ACE2, or both. Cell lines were generated as described in the Materials and Methods. A single dilution of particles was used to infect cells prior to spinoculation as described in the Methods. Infections were done in technical triplicates. Presented are the aggregated results from two independent replicates and error bars show SEM. For statistics, ns = not significant, ** is a p-value <0.01, and **** is a p-value <0.0001. **(B)** Western blot of ACE2 expression in 293T cell lines. This was performed as described in the methods and the values below each column represents the relative abundance of ACE2. **(C)** Normalized CoV2pp entry into single cell clones. Entry was normalized to the wild type parental cell line and further normalized to VSV-G entry. Presented are the average of one experiment in technical triplicates. Error bars show the median and interquartile range. Raw entry data for each cell clone is shown in Supplementary Fig. 9A **(D)** Entry inhibition of CoV2pp in by Nafamostat mesylate, a serine protease inhibitor. Nafamostat was mixed with CoV2pp (left panel) or VSV-Gpp (right panel) prior addition to cells. Shown are the results from one experiment in technical triplicates. Data are presented as described in Fig. 4A and error bars show SEM.

These observations highlight the complex roles that receptor binding and protease activation play in SARS-CoV-2 entry, especially since ACE2 is a known substrate for TMPRSS2,^107^ and TMPRSS2 is also known to undergo autocatalytic cleavage.^108^

Given how TMPRSS2 can enhance ACE2 dependent virus entry in a non-linear fashion, we used BALDpp, CoV2pp, and VSV-Gpp to screen 19 single cell clones derived from 293T-ACE2 or 293T-ACE2+TMPRSS2 or Vero-ACE2 bulk transduced cells. The latter (Fig. 3) served as an additional control in a naturally permissive cell line for SARS-CoV-2 entry and replication. All three bulk transduced cell lines resulted in significant increases in entry of CoV2pp relative to the parental 293T and Vero CCL81 cells (Supplemental Fig. 9 and Fig. 6C). However, only a subset of the single cell clones performed better than bulk transduced cells. This is especially notable in single cell clones derived from 293T-ACE2+TMPRSS2 parentals, where only two of eight single cell clones show greater entry than the bulk transduced cells (Fig. 6C). One particular clone, F8-2 (Fig. 6C) showed a nearly ten-fold increase in CoV2pp entry relative to the bulk transduced cells. Using F8-2 to titer untreated CoV2pp without spinoculation, we observed a dramatic increase in signal:noise relative to Vero-CCL81 WT cells and even the most permissive 293T-ACE2 clone 5-7 (Supplemental Figure 10) such that RLU signals were consistently 100-200 fold over BALDpp even at 1:50 dilution. TMPRSS2 was determined to be the main driver of this entry enhancement in the F8-2 cells as treatment with Nafamostat, a serine protease inhibitor, potently inhibited entry. However, this entry inhibition plateaued at 90% of maximal infection and the remaining 10% is nearly equivalent to the raw RLU values seen with bulk 293Ts stably expressing ACE2 alone (Fig. 6D and Supplemental Figure 9), suggesting a TMPRSS2-independent mechanism of entry. Entry into 293T-ACE2 cells was not inhibited by Nafamostat, once again highlighting that CoV2pp can enter by both the early and late entry pathways that have differential protease requirements.

### Diverse cell lines maintain similar kinetics in CoV2pp viral neutralization assays

We identified sera samples from 15 patients shown in Fig. 4A and tiered them into three groups: negative for CoV2pp neutralization (negative), weakly positive for CoV2pp neutralization (low positive), or strongly positive for CoV2pp neutralization (high positive) (Fig. 7A). We then pooled equal volumes of each set of samples and performed CoV2pp neutralization assays on Vero-CCL81 WT, 293T-ACE2 clone 5-7, 293T-ACE2+TMPRSS2 bulk transduced, and the 293T-ACE2+TMPRSS2 clone F8-2. We demonstrated that even in the case of varying levels of ACE2 and TMPRSS2 expression, CoV2pp neutralization assays show consistent patterns of neutralization, exhibiting the robust nature of the assay in tandem with its sensitivity in detecting relative differences in neutralizing titer (Fig. 7B). Patterns of neutralization as well as the calculated absIC50 and absIC80 reveal a large dynamic range between low and high neutralizing patient sera across cell lines (Fig. 7B).

**Figure 7.**
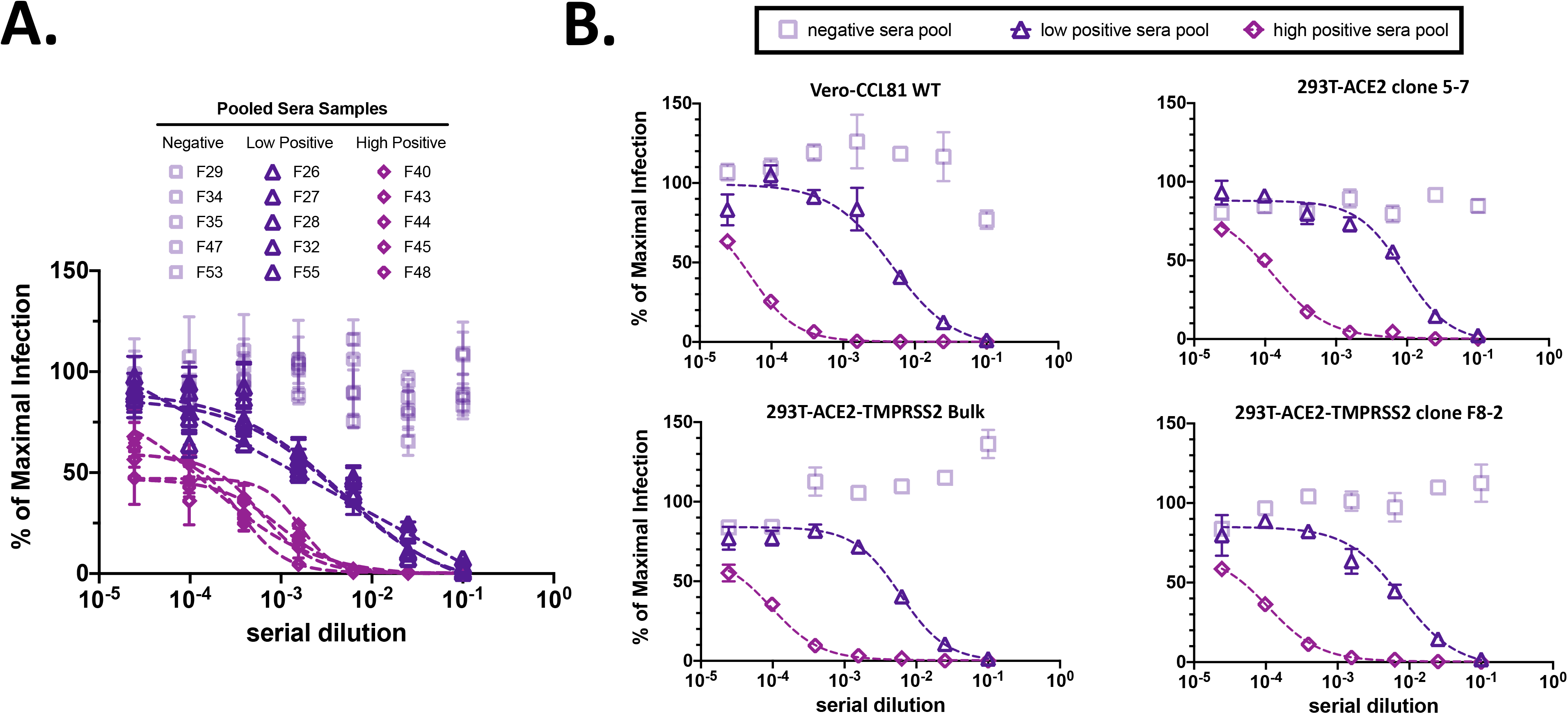
Ultra-permissive 293T-ACE2+TMPRSS2 cell clones retains the same phenotypic sensitivity to convalescent COVID-19 sera. **(A)** Selection of pooled sera samples. Results from Fig. 4A are reproduced here for the reader’s convenience. Presented are the subset of samples that were pooled for use in viral neutralization assays (VNAs) in the adjacent panel. **(B)** Vero CCL81 and transduced 293T cells were used for VNAs. Sera previously shown to be negative, weakly positive, or strongly positive for CoV2pp neutralizations were selected to be pooled in equal volumes. These were subsequently used for VNAs, which were performed and presented as described in Fig. 4A. Notably, these VNAs were performed in the absence of exogenous trypsin or spinoculation.

## Discussion

Here, we present detailed and optimized protocols for producing VSVΔG pseudotyped viral particles bearing SARS-CoV-2 spike protein. These CoV2pp recapitulate the SARS-CoV-2 entry requirement for ACE2 expression on the host cell and enhanced infectivity in the presence of activating proteases such as trypsin and/or TMPRSS2 in both 293T and Vero cells. Evidence from our original standard condition suggested that only a minor fraction of the trypsin added was required, and this trypsin acted at the level of receptor binding on the host cell (Fig. 2C and D). Due to the observed effect of trypsin at the point of infection, we hypothesize that interaction with a cellular factor, likely ACE2, induces conformational changes necessary for further protease-mediated activation, likely at the S2’ cleavage site, of SARS-CoV-2 spike. Moreover, in a competitive inhibition assay, entry by the trypsin-treated CoV2pp was successfully inhibited by sRBD. This faithful recapitulation of the entry processes previously described for SARS-CoV-1 and SARS-CoV-2 suggests that the trypsin treated CoV2pp represent a biologically relevant system for identifying cells that support SARS-CoV-2 entry and for screening for entry inhibitors, especially neutralizing antibodies or patient sera.

Prior to the use of trypsin-treated CoV2pp for neutralization experiments, we assessed how heat inactivation of sera and different cell media affect neutralization. Here, we report detectable neutralization by negative patient sera, which was previously reported in mouse and human sera by Nie et al.^8^ However, it is unclear whether the sera used by Nie et al was heat inactivated. Our observations also raise questions concerning the role the previously mentioned heat-labile serum factor might play *in vivo*. We have shown that the CoV2pp VNA displays high sensitivity to the inhibition of protease-mediated entry enhancement by human serum, FBS, SBTI, and even Nafamostat when the protease in question is TMPRSS2. The inhibitory potential of human serum implies a potential role serum factors could play in SARS-CoV-2 pathogenicity, tissue restriction, and systemic spread in previously SARS-CoV-2 naive patients (manuscript in submission). These findings led to the establishment of heat inactivation of sera and use of DMEM+10% FBS as conditions for trypsin-treated CoV2pp neutralization experiments. When used for viral neutralization assays with patient sera, the absIC50/absIC80 against CoV2pp correlated strongly with full-length spike ELISAs and live virus microneutralization titers. Moreover, we have produced several batches of our CoV2pp and shipped them (along with Vero-CCL81 cells) to many other groups as an “out-of-the-box” neutralization assay. The first three groups to receive these particles, and who have volunteered their data, have successfully screened patient sera with our assay and observed moderate but significant correlations to spike ELISAs.

While ELISAs provide valuable information about epitopes recognized by individual samples and antibody quantities, functional studies allow for more in-depth analyses of neutralization potential. Notably, RBD-binding antibodies, particularly those that can inhibit ACE2 binding, have received a large amount of attention. However, recent studies identifying non-RBD binding—yet still neutralizing—antibodies, lend insight into novel neutralization mechanisms and further highlight the importance of functional neutralization assays.^84,85^ Moreover, our standardized CoV2pp VNA has a large dynamic range that can generate robust neutralization curves, which allows for the calculations of more stringent metrics such as absIC50/absIC80. AbsIC50/absIC80 give a more meaningful description of the neutralization potential of a given serum sample as many patient sera (and potentially vaccine sera) may not even achieve an absIC80. Reporting such standardized metrics will allow more meaningful comparisons of vaccine elicited humoral responses, as well as the neutralization potential of convalescent sera, especially when the latter is used for convalescent plasma therapy. This is of particular importance given the widely variable ratios of spike ELISA binding values and neutralizing antibody titers in comparisons of patients infected by SARS-CoV-2 and patients receiving vaccines for SARS-CoV-2.^13^

Early reports of convalescent sera therapies show a tolerable safety profile and modest benefits from this therapeutic approach. ^17,20,29,31–33,109^ However, many of these trials only consider ELISA neutralization titers and utilize extremely variable ELISA endpoint titers from not reported/available to ranging from 1:40 to >1:1350. Interestingly, one pre-peer reviewed study incorporated functional neutralization studies by utilizing trypsin-treated live virus to screen for sera with >1:80 microneutralization titers on Vero E6 cells.^33^ Given the wide variance in ELISA titers as well as in virus neutralization titers, we believe convalescent plasma therapy will be enhanced if patient sera are functionally screened and limited to only those displaying potent neutralization titers. This will have the benefit of only transfusing patients with convalescent sera that have a strong likelihood of substantial *in vivo* inhibitory potential, which is of particular importance given the volumes transfused relative to a patient’s total blood volume. Given our results, a reasonable threshold might be a VNA-derived reciprocal absIC80 of ≥343.3 (i.e. ≥75th percentile).

Lastly, we utilize the CoV2pp system to screen 19 single cell clones and identify two single cell clones of interest. These clones (293T-ACE2 clone 5-7 and 293T-ACE2+TMPRSS2 clone F8-2) both support effective viral entry in the absence of trypsin and spinoculation and can be used for scaling up viral neutralization assays. The ultra-permissive 293T-ACE2+TMPRSS2 F8-2 clone in particular can support the use of a standardized VNA at the scale needed for screening entry inhibitors, vaccine samples, donor plasma, etc. Our standardized CoV2pp production lot from a single lab at 30x 10-cm dishes was sufficient for ~12,000 infections/week when performed in a 96-well format. The trypsin-treated CoV2pp (diluted 1:4) gives 100:1 signal:noise ratio when performed in a 100 μl infection volume on Vero-CCL81 cells with spinoculation. Using the ultra-permissive F8-2 clone, a 1:50 dilution gives similar signal:noise without any trypsin treatment or spinoculation. Thus, our weekly production lot becomes sufficient now for ~150,000 infections/week, which is enough for generating full neutralization curves for ~4,600 to ~6,200 samples (assuming an 8-point dilution series performed in quadruplicates or triplicates, respectively).

Several recently described systems including VSV encoding the SARS-CoV-2 spike gene^9,110^ and lentiviruses pseudotyped to bear the spike protein,^111^ are capable of serving as surrogate assays for assessing viral neutralization by patient sera or monoclonal antibodies. The replication competent VSV system is attractive but still relies on a truncated spike. We have not been able to rescue one with the full-length tail, even in our F8-2 clone although we could rescue multiple VSVs bearing various betacoronavirus spikes (all with truncated tails). Nonetheless, our standardized CoV2pp based on the VSVΔG system presents many advantages including safety, ease and speed-of-use, identity to full-length SARS-CoV-2 spike, versatility for studying spike mutants, and a large dynamic range. First, the viral genome used in this system lacks a viral glycoprotein, which limits the virus to single-cycle replication and mitigates concerns about viral spread. Next, because of the efficient replication of VSV, this system can be used to further interrogate SARS-CoV-2 entry in primary cells and allows for the detection of Renilla luciferase (or the desired reporter gene) within 12-18 hours post infection. Additionally, the VSVΔG system presented here represents viral entry in the absence of mutations or truncations for enhanced fusogenicity and/or entry dynamics. Lastly, since a viral glycoprotein must be provided in trans for every production, this system is not susceptible to mutations over several passages and is not dependent on repeated, arduous rescue attempts for the study of naturally occurring spike mutants or chimeric spike glycoproteins. These studies may prove beneficial as we consider natural occurring spike mutations—described on platforms such as GISAID—and strive to understand their influence on viral entry kinetics or the influence on escape from antibody neutralization.

In sum, we present detailed and optimized protocols for the production of a BSL-2-safe VSVAG-rLuc pseudoparticle and use it to interrogate viral entry. More importantly, we present several resources that we believe will be invaluable during this global pandemic. This includes cell lines (particularly 293T-ACE2+TMPRSS2 and Vero-CCL81-TMPRSS2 cells), and CoV2pp that are ready to use “out-of-the-box” for mechanistic studies of viral entry or to screen inhibitors of viral entry. Our findings, resources, and proposed guidelines have implications for standardizing viral neutralization assays, with particular importance for screening therapeutic monoclonal antibodies, vaccine efficacy, and convalescent sera.

## Materials and Methods

### Plasmids

⍰ SARS-CoV-2 spike is in a pCAGG backbone and expresses the codon optimized Wuhan-Hu-1 isolate (NCBI ref. seq. NC_045512.2).
⍰ SARS-CoV-2 sRBD (NCBI GenBank MT380724.1 from Krammer lab) is in a pCAGG backbone and expresses the codon optimized sequence from the Wuhan-Hu-1 isolate. sRBD-His used for neutralization studies was generated from this construct.
⍰ VSV-G is in a pCAGG backbone and expresses wild type Indiana strain VSV-G (Genbank: ACK77583.1).
⍰ ACE2 packaging construct (GeneCopoeia, cat no EX-U1285-Lv105) uses a CMV promoter to express TMPRSS2 and bears a puromycin selection marker in the integrating cassette.
⍰ TMPRSS2 packaging construct (GeneCopoeia, cat no EX-Z7591-Lv197) uses a CMV promoter to express TMPRSS2 and bears a blasticidin selection marker in the integrating cassette.
⍰ psPAX2 2^nd^ generation lentiviral packaging plasmid (Addgene #12259) expresses HIV-1 Gag, Pol, and Pro proteins.
⍰ NiV-RBP is in a pCAGG backbone and expresses the HA-tagged codon optimized NiV receptor binding protein.

All plasmids listed here are ampicillin resistant. These constructs were transformed into stellar competent cells, grown in bacterial growth media containing carbenicillin, prepared using Invitrogen’s midiprep kit, and sequence verified prior to use for experiments.

### Maintenance and generation of cell lines

Vero-CCL81 and 293T cells were cultured in DMEM with 10% heat inactivated FBS at 37°C with 5% CO_2_. VSV-G pseudotyped lentiviruses packaging ACE2 or TMPRSS2 expression constructs were generated by using Bio-T (Bioland; B01-01) to transfect 293T cells with the second-generation lentiviral packaging plasmid (Addgene; 12259), pCAGG-VSV-G, and the desired expression construct (i.e. ACE2 or TMPRSS2). The media was changed the next morning. Viral supernatant was collected 48 hours post transfection, clarified by centrifugation at 4000 rpm for 5mins, and aliquoted prior to storage at −80°C. Vero-CCL81 and 293T cells were transduced in a 6-well plate with the prepared lentiviral constructs. Two days after transduction, these cells were expanded into a 10cm plate and placed under selection with puromycin (for ACE2 transduced cells) or blasticidin (for TMPRSS2 transduced cells). 293T and Vero-CCL81 cells were selected with 2 or 10μg/mL of puromycin, respectively. For blasticidin, 293T were selected with 5μg/mL and Vero-CCL81 cells were selected with 15μg/ml. To generate ACE2 and TMPRSS2 expressing 293T cells, 293T-ACE2 cells were transduced with the VSV-G pseudotyped lentivirus packaging TMPRSS2. These cells were subsequently selected with 5^g/mL blasticidin. Low passage stock of each cell line generated were immediately frozen down using BamBanker (Fisher Scientific; NC9582225). Single cell, isogenic clones were isolated via serial dilution in a 96 well plate. Wells with only a single cell were grown up and eventually expanded while under selection.

### Pseudovirus production and titering

We provide detailed production and titering protocols in the supplementary text (Supplementary Methods). Briefly, 293T producer cells were transfected to overexpress SARS-CoV-2 or VSV-G glycoproteins. For background entry with particles lacking a viral surface glycoprotein, pCAGG empty vector was transfected into 293T cells. Approximately 8 hours post transfection, cells were infected with the VSVΔG-rLuc reporter virus for 2 hours, then washed with Dulbecco’s phosphate-buffered saline (DPBS). Two days post infection, supernatants were collected and clarified by centrifugation at 1250 rpm for 5mins. Upon collection, a small batch of VSVΔG-rLuc particles bearing the CoV2pp were then treated with TPCK-treated trypsin (Sigma-Aldrich; T1426-1G) at room temperature for 15 minutes prior to inhibition with soybean trypsin inhibitor (SBTI) (Fisher Scientific; 17075029). Particles were aliquoted prior to storage in −80°C to avoid multiple freeze-thaws.

To titer these pseudoviruses, 20,000 Vero-CCL81 cells were seeded in a 96 well plate 20-24hrs prior to infection. A single aliquot of BALDpp, CoV2pp, and VSV-Gpp were used for infections and titrations were performed in technical triplicates. At 18-22 hours post infection, the infected cells were washed with DPBS, lysed with passive lysis buffer, and processed for detection of Renilla luciferase. The Cytation3 (BioTek) was used to read luminescence. Additional details can be found in Supplementary Methods.

### Collection of producer cells and concentration of pseudotyped particles

Cell lysates were collected from producer cells with 10mM EDTA in DPBS. Cells were subsequently lysed with radioimmunoprecipitation assay (RIPA) buffer (Thermo Scientific, 89900) containing protease inhibitor (Thermo Scientific, 87785) for 30 minutes on ice. Lysates were centrifuged at 25,000 × g for 30 minutes at 4°C, and the supernatants were collected and stored at −80°C. Total protein concentrations were determined by the Bradford assay. For viral pseudoparticles, 10 mL of designated viral particles was concentrated via 20% sucrose cushion (20% sucrose in DPBS), Amicon Ultra centrifugal filter (100 kDa cutoff, Millipore Sigma, UFC910024), or PEG precipitation (Abcam, ab102538). Concentrated viral particles were resuspended in 300μL of PBS or Opti-MEM for further analysis.

### Western blots

All protein samples were run under reduced conditions by dilution in 6X SDS containing dithiothreitol (DTT) and 5% beta-mercaptoethanol (Fisher Scientific; ICN19483425). The protein was subsequently incubated in a heating block at 95°C for 15mins, run on a 4-15% SDS-PAGE gel, and transferred to polyvinylidene difluoride (PVDF) membranes (Bio-Rad). Membranes were blocked with phosphate-buffered saline blocking buffer (LI-COR, 927-700001), and then probed with the indicated antibodies. Antibodies against SARS-CoV2 (2B3E5 from Dr. Thomas Moran and GTX632604 from GeneTex), ACE2 (66699-1-Ig from Proteintech and Rb ab108252 from abcam), VSV-G (A00199 from Genescript), VSV-M (EB0011 from Kerafast), anti-HA (NB600-363 from Novus), and CoX IV (926-42214 from LI-COR) were used. For secondary staining, membranes were washed and incubated with the appropriate Alexa Fluor 647-conjugated anti-mouse antibody or Alexa Fluor 647-conjugated anti-rabbit antibody. Alexa Fluor 647 was detected using the ChemiDoc MP imaging system (Bio-Rad). Relative ACE2 or TMPRSS2 abundance was calculated by first normalizing abundance relative to GAPDH expression, then normalizing to wild type expression.

### RNA extraction and qPCR for ACE2 and TMPRSS2 expression

Total RNA was extracted from cells using Direct-zol^TM^ RNA Miniprep kit (Zymol, R2051), and reverse transcription (RT) was performed with the Tetro™ cDNA Synthesis kit (Bioline, BIO- 65043) and random hexamers. RT PCR was performed with the SensiFAST™ SYBR & Fluorescein Kit (Bioline, BIO-96005). For qPCRs, HPRT forward (5’-ATTGTAATGACCAGTCAACAGGG-3’) and reverse (5’-GCATTGTTTTGCCAGTGTCAA-3’) primers, ACE2 forward (5’-GGCCGAGAAGTTCTTTGTATCT-3’) and reverse (5’-CCCAACTATCTCTCGCTTCATC-3’) primers, and TMPRSS2 forward (5’- CCATGGATACCAACCGGAAA-3’) and reverse (5’-GGATGAAGTTTGGTCCGTAGAG-3’) primers were utilized. Samples were read on the CFX96 Touch Real-Time PCR Detection System (Biorad). For qPCR forward and reverse primers were utilized. The qPCR was performed in duplicates for each sample and results were calculated using 2^-ΔΔCT^ with normalization to the HPRT housekeeping gene control and further normalization to the 293T parental cells.

### Sera acquisition

All patient sera were acquired after approval by the respective institutional review boards and/or equivalent oversight bodies (Bioethics Committee, Independent Ethics Committee) are indicated: (1) Mount Sinai Hospital Institutional Review Board (New York, USA), (2) Louisiana State University Health Sciences Center – Shreveport (LSUHS, Louisiana, USA), and (3) Fundacion Instituto Leloir-CONICET, Universidad Nacional de San Martin, Laboratorio Lemos SRL, Universidad de Buenos Aires (COVIDAR Argentina Consortium, Buenos Aires, Argentina). Samples were de-identified at the source institutions or by the respective PIs of the IRB approved protocols for sample collection before analysis performed in this study. All necessary patient/participant consent has been obtained and the appropriate institutional forms have been archived.

### ELISAs and Live Virus Neutralization

Spike ELISAs for patient sera from the Krammer lab were performed in a clinical setting using the two-step protocol previously published (Mount Sinai Hospital). Briefly, this involves screening patient sera (at a 1:50 dilution) with sRBD and samples determined to be positive were further screened at 5 dilutions for reactivity to spike ectodomain. All 36 samples were screened in this manner, but a subset of 15 samples were further screened for IgG and IgM binding antibodies to spike ectodomain. The protocol from Stadlbauer et al^106^ was modified slightly to start from a 1:300 and end at a 1:24300 dilution of sera. IgG and IgM antibodies were detected with secondary antibodies conjugated to HRP (Millipore AP101P for anti-Human IgG and Invitrogen A18841 for anti-Human IgM). Background was subtracted from the OD values, samples were determined to be positive if ≥ 3 fold over the negative control and AUC was calculated in PRISM. ELISAs performed by the LSUHS group utilized sRBD with a 1:50 dilution of patient sera to screen all samples followed by spike ectodomain with patient sera at a 1:100 dilution. Background subtracted OD values are reported for both sets of ELISAs. ELISAs performed by the COVIDAR group utilized a mixture of sRBD and spike ectodomain for samples serially diluted from 1:50 to 1:6400. AUC were calculated as described above.

All live virus neutralizations were performed at biosafety-level-3 (BSL-3) using the USA-WA/2020 isolate of SARS-CoV-2 as described in Amanat et al^73^. Briefly, ~600 50% tissue culture infectious doses (TCID50) of virus was incubated with a serial dilution of patient sera for 1hr at 37°C prior to infection of Vero-E6 cells. Forty-eight hours post infection, cells were fixed in 10% PFA and stained with mouse anti-SARS-CoV nucleoprotein antibody. This was subsequently detected by the addition of HRP-conjugated goat anti-mouse IgG and SIGMAFAST OPD. The BioTek Synergy 4 plate reader was used to measure OD490, which was subsequently used to calculate microneutralization (MN) titers. The samples with live virus MN titers were a part of a larger study by Krammer and colleagues looking at the longitudinal dynamics of the humoral immune response. This study was recently posted on medRxiv.^35^ We obtained permission from the authors to utilize a random subset of sera samples from their study and their associated MN titers for validation studies with our CoV2pp based virus neutralization assay.

### Neutralization studies with patient sera, soluble RBD, or Nafamostat-mesylate

De-identified sera were obtained with IRB approval to use for research purposes. Unless otherwise noted, all patient sera were heat inactivated at 56°C for 30 minutes, and serially diluted in DMEM+10%FCS when performing virus neutralization assays (VNAs). For groups receiving our CoV2pp, we recommended titrating our stocks first to determine the linear dynamic range that would be useful for VNAs done in their labs. As a quality control, we only send out CoV2pp stocks that give signal:noise ratios of at least 100-fold over BALDpp when diluted 4-fold in 100 μl total infection volume in 96-well plate format. For the VNAs performed in our lab (ISMMS-1), a pre-titrated amount of pseudotyped particles (diluted to give approximately 10^5^ RLU) was incubated with a 4-fold serial dilution of patient sera for 30 minutes at room temperature prior to infection of Vero-CCL81 cells seeded the previous day. For sRBD or Nafamostat inhibition, a pre-titrated amount of pseudotyped particle dilution was mixed with the protein or compound and added to cells immediately after. Approximately 20 hours post infection, cells were processed for detection of luciferase activity as described above. Our recommendations to generate a robust neutralization curve were to do an 8-point serial dilution curve with each point done in triplicate. Raw luminometry data were obtained from labs that volunteered VNA results from at least 12 patient samples and analyzed as indicated below.

Method modifications from the three contributing labs are as follows. Serum neutralizations by LSUHS (Kamil and Ivanov) were performed by first diluting 4-fold in 100 μl total volume then diluting via a 3-fold serial dilution. Cell lysates were transferred to a white walled 96 well plate, then the Promega Renilla luciferase assay kit was utilized to detect luciferase. Plates were read on a Tecan SPARK plate reader by collecting total luminescence signal for 10 seconds. ISMMS-2 (Hioe) began neutralizations at a 10-fold dilution and proceeded with a 4-fold serial dilution. Plates were read on a black walled 96 well plate using the Renilla Glo substrate (Promega, E2720) with a 1 second signal integration time. COVID-19 samples were provided to ISMMS-2 by the Clinical Pathology Laboratory at ISMMS or from an IRB-approved study at the James J. Peters VA Medical Center. COVIDAR (Gamarnik) began at either an 8-fold or 16-fold dilution then continuing with either a 3-fold or 2-fold serial dilution respectively. White, F-bottom Lumitrac plates (Greiner, 655074) plates were read via the GloMax® Navigator Microplate Luminometer (Promega, GM200) using the ONE-Glo™ Luciferase Assay System (Promega, E6110).

### Inhibitory Concentration Calculations and other R packages used

Relative inhibitory concentrations (IC) values were calculated for all patient sera samples by modeling a 4-parameter logistic regression with drm in the R drc package.^112^ For examples, a relative inhibitory concentration of 50% (IC50) is calculated as the midway point between the upper and lower plateaus of the curve. Absolute inhibitory concentration (absIC) was calculated as the corresponding point between the 0% and 100% assay controls. For example, the absIC50 would be the point at which the curve matches inhibition equal to exactly 50% of the 100% assay control relative to the assay minimum (0%).^113^ As a result, sera samples that are non-neutralizing or minimally neutralizing may have lower plateaus indicating they cannot reach certain absolute inhibitory concentrations, such as an absIC90 or absIC99. R was also used to generate the heatmaps presented in Fig. 4B and Fig. 5A as well as the plots in Fig. 4D, Fig. 5C, and Supplemental Fig. 7.

## Data Availability

The authors confirm that the data supporting the findings of this study are available within the article and/or its supplementary materials.

## Acknowledgements

In addition to the three labs that provided data for this paper, we acknowledge the generosity of the many labs that independently verified our CoV2pp VNA. We want to especially acknowledge the entirety of the Lee Lab, whom, during a pandemic, dropped all other work and pooled resources together in pursuit of developing this and other tools that are providing support to other labs doing COVID-19 work. Moreover, we acknowledge the generosity of the patients that donated blood for use in convalescent sera studies and for further use in research studies similar to the work presented here.

The authors acknowledge the following funding: KYO and CS were supported by Viral-Host Pathogenesis Training Grant T32 AI07647; KYO was additionally supported by F31 AI154739. SI and CTH were supported by postdoctoral fellowships from CHOT-SG (Fukuoka University, Japan) and the Ministry of Science and Technology (MOST, Taiwan), respectively. BL acknowledges flexible funding support from NIH grants R01 AI123449, R21 AI1498033, and the Department of Microbiology and the Ward-Coleman estate for endowing the Ward-Coleman Chairs at the ISMMS. JPK and SSI acknowledge funding from a LSUHS COVID-19 intramural grant. JPK and SSI acknowledge additional funding from NIH grants AI116851 and AI143839, respectively. This work was further supported by the Microbiology Laboratory Clinical Services at the Mount Sinai Health System and the Mount Sinai Health System Translational Science Hub, NIH grant U54TR001433 to the Department of Medicine of the ISMMS to SZP. We also acknowledge funding from the Department of Veterans Affairs Merit Review Grant I01BX003860 (CEH, SZP, and JK), Research Career Scientist Award 1IK6BX004607 (CEH), and NIH grant AI139290 (CEH, SZP). We thank Randy A. Albrecht for oversight of the conventional BSL3 biocontainment facility. Work in the Krammer laboratory was partially supported by the NIAID Centers of Excellence for Influenza Research and Surveillance (CEIRS) contract HHSN272201400008C (FK), Collaborative Influenza Vaccine Innovation Centers (CIVIC) contract 75N93019C00051 (FK) and the generous support of the JPB foundation, the Open Philanthropy Project (#2020-215611) and other philanthropic donations.

---

**Supplemental Table 1. Descriptive statistics for CoV2pp neutralizations across 4 groups**. Presented are the descriptive statistics from the CoV2pp neutralizations shown in Fig. 5C. Absolute IC50, 80, and 90 were calculated as previously described in the Methods. Median and other percentiles presented here were calculated in PRISM.

**Supplemental Figure 1. Expression spike glycoproteins in different growth media**. Expression of CoV-2 spike in producer cells shows modestly increased cleavage in the presence of reduced or absent FBS. Western blots performed as described in the Methods.

**Supplemental Figure 2. Dilution of CoV2pp in the absence of serum free media produces the highest signal:noise for trypsin treated CoV2pp**. Performed as described in Fig. 2B, but was done in the absence of spinoculation. Presented are the results from an experiment in technical triplicate and error bars show the SEM.

**Supplemental Figure 3. Sera neutralization in the absence of 10% FBS and optimization of neutralizations**.

**(A)** Negative sera potently inhibits trypsin treated CoV2pp. CoV2pp were diluted in serum free media (SFM), then pooled negative sera and a positive serum were used to neutralize entry. An aliquot was heat inactivated (HI) for 1hr in a 56°C water bath prior to use. Neutralizations were performed as described in the methods. Data are presented on a linear (left panel) and log scale (right panel). Each replicate from one experiment in technical duplicates are shown and neutralization curves were generated as done in Fig. 1D. **(B)** Sera neutralizations were performed with untreated CoV2pp (left panel) or CoV2pp treated with trypsin (middle panel). Both particles were diluted in DMEM+10% FBS and neutralization curves are presented as described above. VSV-G was not neutralized by the negative or positive sera (right panel). **(C)** sRBD neutralizes CoV2pp equivalently across all conditions tested. Data presented in Fig. 1D (i.e. the untreated CoV2pp) is duplicated here. Neutralization curves are presented as described above.

**Supplemental Figure 4. Live SARS-CoV-2 microneutralization curves**. Live virus microneutralization curves were performed as described in the materials and methods. Neutralizations were performed in technical duplicates and shown are SD. Data are presented as in Fig. 4A and fit to a variable slope, 4-parameter logistics curve.

**Supplemental Figure 5. Neutralization curves from the LSUHS, ISMMS-2, and COVIDAR labs**. The neutralization curves presented here were generated from the same data used to create the neutralization heat maps shown in Figure 5A. The curves were fit using a variable slope, 4 parameter logistics model (robust regressions fitting). The ISMMS-2 group (top left panel) and COVIDAR group (bottom panels) perform neutralizations in technical triplicates. The LSUHS group (top right panel) performed their neutralizations in technical quadruplicates.

**Supplemental Figure 6. Relationship of ELISA endpoint titers and CoV2pp reciprocal AbsIC50, 80, and 90**. Presented are the clinical lab ELISA endpoint titers and CoV2pp neutralization absIC values. There are 11 samples with ELISA endpoints of 320, 8 samples with ELISA endpoints of 960, and 11 samples with ELISA endpoints of 2880. Absolute (Abs) IC50, 80, and 90 were calculated as described in the Methods. Error bars (blue) show median and interquartile range, red and black dotted lines represent the median and 75^th^ percentile for each AbsIC value as calculated in Supplemental table 1. The gray shaded region indicates samples that fall above the 75^th^ percentile. One sample with an ELISA endpoint of 320 has an absIC90 below 10^-1^ and thus is not present on the absIC90 graph.

**Supplemental Figure 7. Ordered CoV2pp absolute IC50 (top), IC80 (middle) and IC90 (bottom) plots from all four groups**. As previously presented in Fig. 4D, the IC50 graph is colored and ordered to display samples with low, average or high IC50 as blue, grey or red circles, respectively. The colors from the IC50 graphs are retained in the IC80 and IC90 graphs, which are ordered from lowest to highest neutralization. Tukey box and whisker plots are presented to the right of the graph. These show findings that are consistent to the observations in Fig. 4D, suggesting that not all samples with high IC50s have potent IC80s or IC90s.

**Supplemental Figure 8. Comparison of CoV2pp Absolute IC values across all 4 groups**. CoV2pp Absolute IC values were calculated as previously described in Fig. 4C. Shown are the CoV2pp absolute IC50 (left panel), IC80 (middle panel) and IC90 (right panel) from all four groups with error bars (blue) showing the median and interquartile range. The red dotted line presents the median from the aggregated positive neutralization samples as reported in Supplemental Table 1. The black dashed line indicates neat serum and the shaded gray region highlights samples that fall below this value. An ordinary one-way ANOVA with Dunnett’s correction for multiple comparisons was performed for statistics. This analysis revealed no statistically significant difference between the Absolute IC values obtained across the 4 groups. There were notable outliers in this data set, including individuals that show poor neutralization (i.e. the 3 samples in the IC50 plot from LSUHS) and an individual that showed exceptionally potent neutralization (i.e. the sample in all plots from ISMMS-2). One sample from ISMMS-1 had an absIC90 below 10^-1^ and, as a result, is not presented on the absIC90 graph.

**Supplemental Figure 9. Screening and validation of single cell clones**. (A) Raw RLU values from infection of the indicated cells by BALDpp, CoV2pp, or VSV-Gpp. Parental cell lines, bulk transduced cell lines, and isogenic cell lines are indicated. Highlighted in blue and purple are the ultra-permissive clones stably expressing ACE2 or ACE2 and TMPRSS2, respectively. Presented are the results from an experiment in technical triplicates and error bars show the SEM. (B) Expression of ACE2 and TMPRSS2 in select cell lines. RNA extraction and qPCR performed as described in the Methods prior to calculating 2^-ΔΔCT^, which was then normalized to the 293T parental cells. Of interest are the clones highlighted in blue and purple, which were transduced to stably express ACE2 or ACE2 and TMPRSS2.

**Supplemental Figure 10. CoV2pp were titered on Vero-CCL81 cells, 293T-ACE2 clone 5-7, and 293T-ACE2-TMPRSS2 clone F8-2**. Titrations were performed with untreated CoV2pp and without spinoculation. Presented are the results from technical triplicates and bars show the SEM.

